# The effect of maternal choline intake on adolescent offspring cognition in adolescence: protocol for a 14-year follow-up of a controlled feeding trial

**DOI:** 10.1101/2025.02.27.25322999

**Authors:** S.A Roth, A. Lam, B.J. Strupp, R.L. Canfield, E.A Larson

## Abstract

**Background:** Choline is an essential micronutrient crucial for fetal neurodevelopment. Numerous rodent studies reveal that maternal prenatal choline deficiency produces lifelong offspring cognitive deficits and that maternal choline supplementation produces lifelong improvements to offspring cognition. Few studies have evaluated this question in humans, and with mixed results.

Nonetheless, the available data raise concerns about the low choline intakes of pregnant women and highlight the need for knowledge on the functional consequences of various choline intakes during pregnancy. To address this, the present study evaluates the cognitive and affective functioning of adolescents born to women who participated in a randomized controlled trial of two levels of choline intake during pregnancy.

**Methods:** In a double-blind controlled choline feeding trial, (N = 26) third-trimester pregnant women were randomly assigned to daily choline consumption at 480 mg/d or 930 mg/d. In this 14-year follow-up, the offspring (n = 21) of these women will complete cognitive tests proctored over conferencing software. We will also assess facets of mental health using the Achenbach System of Empirically Based Assessment. These assessments will test the hypothesis that third trimester maternal choline intake exerts lasting effects on offspring attention, memory, executive function, and mental health.

**Significance and Impact:** We hypothesize that adolescent offspring born to women in the 930 mg/d group will perform better in domains of memory, attention, executive function, and mental health than offspring of the 480 mg/d group. This study is unique because total maternal choline intake is precisely known, and the offspring are followed into adolescence, a time when group differences are indicative of lifelong effects of the prenatal intervention. The findings will provide important new information concerning the lasting functional consequences of maternal choline intake during pregnancy for offspring neurobehavioral health, thereby informing dietary recommendations and supplementation policies for pregnant women.

**Trial registration:** ClinicalTrials.gov: NCT05859126

## Introduction

Choline is an essential micronutrient that is critical for fetal neurodevelopment (1). Extensive rodent research has consistently demonstrated the importance of maternal choline intake for offspring neurodevelopment, cognition, and brain health into old age (1-5). Specifically, maternal choline deficiency in rodents produces lasting offspring cognitive deficits (2–4). Conversely, increasing the mother’s choline intake to approximately 4.5 times the amount contained in standard rodent chow improves offspring memory and attention (3–6). In addition, increased maternal choline intake offers protection against a variety of neurotoxic agents (e.g., alcohol, manganese) as well as against neurological disorders (e.g., Alzheimer’s Disease, Down syndrome, Autism; (2,3,7–14). Preliminary evidence in rodents also suggests that maternal choline intake may play a role in the offspring’s vulnerability to stress, anxiety, and depression (15,16).

In contrast to the robust rodent literature, relatively few studies have assessed the impact of varied maternal choline intakes on offspring cognition in humans. The available observational and experimental studies are discussed below.

The majority of human studies evaluating the impact of maternal choline intake on offspring cognition have been observational. Three such studies report significant associations between low maternal choline intake or plasma choline and poorer offspring cognition (17–19), but two similar studies find no such association (11,20). Additionally, three studies point to a potential neuroprotective effect of higher maternal plasma choline against common early insults, including viral infections and cannabis exposure (21–23). One factor which may have contributed to these mixed results is that these studies have necessarily relied on either self-reported dietary intake or blood biomarkers of choline status, both of which are imprecise, potentially leading to exposure misclassification. Importantly, too, all these observational studies are limited by the possibility of uncontrolled confounding, which hinders the ability to infer causation.

Many of the limitations of these observational studies can be overcome with a randomized controlled trial (RCT) design. However, few human studies have experimentally manipulated maternal choline intake during pregnancy and assessed offspring cognition (24–26). One such study evaluated the effect of providing pregnant women with either 900 mg/d choline supplement or placebo from week 17 of gestation through birth and found a beneficial effect of the choline supplement on 5-month-old infants’ auditory attentional gating (17). Later, when these same offspring reached 40 months of age, a parental report instrument indicated that the offspring of supplemented mothers had fewer attentional problems and fewer withdrawn behaviors than those born to women in the placebo group (26). However, the study did not include direct behavioral measures of offspring cognition or behavior. A second study randomized pregnant women to receive either 750 mg/d choline supplement or placebo from 18 weeks of gestation through 90 days postpartum and found no significant group differences in infants’ short-term visuospatial memory or long-term episodic memory at either 10 or 12 months of age (24). In both studies, there were methodological concerns, including post-hoc alterations to planned analyses and uncertain adherence to the assigned choline or placebo dosing regimen (24,26).

The third experimental study evaluating the effect of maternal choline intake on offspring cognition was conducted in our lab. This study was a double-blind randomized controlled feeding trial, in which total maternal choline intake was controlled at one of two levels (480 vs. 930 mg choline/d) throughout the third trimester of pregnancy (25,27). Cognitive testing at four time points during the first year of life revealed a beneficial effect of the higher maternal choline intake on offspring information processing speed (16). Subsequent testing of these same children at age 7 years revealed the long-term benefits of the higher maternal choline intake on offspring attention, working memory, and executive functioning (25,28).

Notably, no RCTs have evaluated the effects of choline intake during pregnancy on offspring cognition during adolescence, a life stage when cognitive performance can better indicate the enduring effects of maternal choline intake (25,29). To fill this knowledge gap, we designed the present study to assess cognitive functioning and mental health in the 14-year-old offspring of the women who participated in the controlled feeding trial mentioned above. In this follow-up study, we will assess cognitive functioning using tests of attention, memory, and executive functioning, domains of cognition shown to be affected by maternal choline intake in animal studies (2–5). In addition, we will assess multiple aspects of adolescent behavior and mental health using a standardized self-report questionnaire designed for adolescents (2–5).

Findings from this study will provide important information about the long-lasting functional consequences of maternal prenatal choline intake on offspring cognition. Choline is not currently included in standard prenatal vitamin regimens, and little is known about the sequelae of the generally low choline intakes seen in women of reproductive age, 90% of whom consume less than the current Adequate Intake (AI) level (30). As such, this work may have important implications for perinatal care and may help elucidate the importance of maternal choline intake for enduring offspring health.

## Methods/Design

### Study Design

The original study was a randomized, double-blind, controlled choline feeding study. Pregnant women entering their third trimester (N = 26) were randomly assigned to consume either 480 mg choline/d (roughly the AI level) or 930 mg choline/d (roughly double the AI level) (27). All participants consumed the same, study-provided, standardized diet containing 380 mg/d of choline. In addition to the standardized diet, participants received a daily choline chloride supplement of either 100 mg or 550 mg, mixed in a cranberry-grape drink, which brought their total daily choline intakes to either 480 mg/d or 930 mg/d. On weekdays, participants consumed at least one of their meals and the choline drink under researcher supervision (27). Participants received all other meals and their weekend daily supplements in take-out containers for consumption at home (27).

### Recruitment

Between January 2009 and October 2010, N= 26 third-trimester pregnant women from the Ithaca, NY, USA area enrolled in the original controlled feeding study (25). At enrollment, the mothers had an average age of 29 years, and participants’ self-reported ethnicities reflected the racial and ethnic distribution of the Ithaca region (25).

For the present follow-up study, our population included the 14-year-old offspring who were born to the women from our controlled feeding study. To recruit, we first contacted participants via their last known email in our participant database. If no email was available, we used their last known phone number. If no contact information was available or if the email/phone number was no longer in use, we utilized web searches to find updated contact information. If participants were non-responsive after two emails and/or three phone calls, we considered them lost to follow-up and noted them as such in the participant tracking log.

Inclusion in this follow-up study required the adolescents to be proficient in English (B2 level equivalent) and to be 14 years old during the testing period. We excluded participants if they had developed a new auditory or visual disability since the last testing period at age 7 or if they were unable to access the internet during the testing period.

Of the original 26 participants, we successfully enrolled 21 offspring who reside in the USA, China, Scotland, and India for the 14-year remote follow-up. Recruitment started in August of 2023 and was concluded in October of 2023.

### Study Status

Data collection is ongoing and will be completed by Spring 2025.

### Behavioral Assessments and Hypothesized Effects

We are conducting remote cognitive and mental health assessments of the teenage offspring to assess the long-term impacts of maternal prenatal choline intake. Remote assessment will allow us to maximize participant retention, as many of our original participants no longer reside in the Ithaca, NY, USA area. We are assessing adolescent cognition, in the domains of memory, attention, and executive functions, using web-based software from the Cambridge Neuropsychological Test Automated Battery (CANTAB Connect). We are also assessing adolescent mental health using the Achenbach System of Empirically Based Assessment (ASEBA) online platform to administer the Youth Self Report (YSR) (31).

Below we describe the tasks and primary outcomes pertaining to each assessment domain (attention, memory, executive functioning, mental health), together with the hypothesized results—stated as a comparison of the 930 mg/d group in relation to the 480 mg/d group.

### Attention

We are assessing the domain of attention using the CANTAB sustained attention task, Rapid Visual Information Processing (RVP). In RVP, a series of individual digits from 2-9 are briefly displayed in a white box at the center of the iPad screen. The digits appear at a rate of 100 digits per minute for 7 minutes. The participant must identify every instance when a given target sequence of 3 digits (e.g., 3-2-5) has appeared in the stream of numbers by pressing a central button as quickly as possible. Superior attention is indicated by the ability to correctly identify target sequences while avoiding false alarms (i.e., responses to nontarget sequences), as summarized by the signal detection parameter A-Prime (32). We hypothesize that participants from the 930 mg/d group will have greater A-Prime scores. A-prime measures the subject’s sensitivity for detecting the three number target sequence, corrected for biased responding. The expected range of A prime scores is 0.00 to 1.00, with 1.00 indicative of excellent sustained attention—a participant that identifies every target sequence throughout the 7-minute test without committing any false alarms.

### Memory

We will use four CANTAB tasks to assess visual and spatial, short-term and long-term memory. These tasks are the Paired Associates learning task, the Delayed Matching to Sample task, and the Spatial Span task, and the Pattern Recognition Memory task.

<u>The CANTAB Paired Associates Learning task (PAL)</u> assesses visual and spatial short-term memory by requiring the participant to remember the location in which they had previously seen a specific pattern appear. For this task, 12 boxes are displayed around the perimeter of the screen. In a pseudo-random order, each box opens briefly to reveal it contains either a unique pattern or nothing. The number of patterns hidden on given trial increments from 2 to 4, 6, 8, and then 12. Memory is tested in each trial by displaying each previously seen pattern in the center of the screen and then asking the participant to select the box location where that pattern was originally located. The number of selection errors is recorded. Here, we hypothesize that participants from the 930 mg/d group will have superior visual and spatial short-term memory, as indicated by fewer errors made when selecting the location of the box where the pattern had originally been located.

<u>Delayed Matching to Sample (DMS)</u> assesses visual matching ability and short-term visual memory for non-verbalizable patterns. Trials of DMS begin by displaying a single target pattern, followed by four similar patterns in a row below the single pattern, one of which matches the target pattern. For trials testing simultaneous visual matching ability, the target pattern remains visible while the participant seeks the matching pattern from among the set of four. For trials testing visual memory, the target pattern disappears and then the four choice patterns are displayed after a delay of either 0, 4, or 12 seconds. The number of correct matches is recorded for each trial type. We hypothesize that participants from the 930 mg/d group will have superior short-term visual memory, as indicated by a greater proportion of correct matches after a delay.

<u>The Spatial Span (SSP)</u> task measures short-term spatial memory capacity. Each SSP trial begins with 9 boxes displayed on the screen. A subset of squares briefly change color in a pseudo-random sequence, and the participant must then touch the boxes that changed color in the same order in which they were displayed. The sequence of boxes that change color increases from two to a maximum of nine, with participants allowed three attempts to correctly reproduce a sequence at each level. The primary endpoint is the longest sequence of boxes touched in the correct order on at least one trial (referred to as span length). We hypothesize that participants from the 930 mg/d group will have a greater span length, indicating superior short-term spatial memory.

<u>The Pattern Recognition Memory (PRM)</u> task assesses short-term and long-term memory for visual patterns. The task displays a series of visual patterns, one at a time for two seconds each, in the center of the screen. This is followed by forced-choice memory test trials in which the participant must discriminate the previously displayed pattern from a similar but novel pattern. For the test of short-term memory, the test trials begin immediately after the patterns are displayed. For the test of long-term memory, the test trials occur following a 20-minute, distraction-filled delay. We hypothesize that participants from the 930 mg/d group will have superior long-term visual recognition memory, as indicated by a greater proportion of correctly recognized patterns after the 20-minute delay.

### Executive Function

To assess our third aim of evaluating group differences in executive functioning, we will focus on the component processes of planning, working memory, inhibitory control, and attentional flexibility. These functions will be assessed using four CANTAB tasks, the Stockings of Cambridge, the Spatial Working Memory task, the Cambridge Gambling Task, and the Intra- Extradimensional Shift task.

<u>Stockings of Cambridge</u> assesses the planning area of executive functioning. In the task, there are two displays, one at the top and one at the bottom of the screen. Each display contains three ‘stockings’ filled with differently colored balls. The balls are arranged in different patterns in each display, and the participant must move the balls in the bottom display to match the pattern shown in the top display. The balls can only be moved one at a time, and the participants are instructed to try and solve the task using the minimum number of moves (referred to as a perfect solution). We hypothesize that the 930 mg/d group will exhibit superior planning ability as indicated by a greater number of perfect solutions.

<u>The Spatial Working Memory</u> test assesses the working memory component of executive functioning. Colored boxes are depicted across the screen. By selecting boxes and using a process of elimination, the participant must find one yellow token hidden within a box. One token is hidden at a time, and a token is not hidden in the same box twice. We hypothesize that the 930 mg/d group will commit fewer total errors in the Spatial Working Memory test, indicative of superior working memory.

<u>The Cambridge Gambling Task</u> assesses risk-taking behavior, which provides an index of inhibitory control. The task displays a row of boxes at the top of the screen– some boxes are red, and others are blue. The ratio of red and blue boxes varies, but one box always contains a hidden token. The participant must select ‘Red’ or ‘Blue’ to choose the box color where they think the token is hidden. Then, participants must decide how much they want to bet on their decision.

The participant gains or loses bet points, depending on the correctness of their decision, given the token location. We hypothesize that the 930 mg/d group will attain a smaller delay aversion score, indicative of better impulse control.

<u>Intra-Extra Dimensional Set Shift</u> assesses rule acquisition and reversal. In this task, colored shapes and white lines appear on screen. These act as two sources of stimuli, simple and complex. The participant is presented with the simple and complex stimuli in pairs, and the participant must determine a preset matching rule and select the correct pair. The participant uses feedback to select the correct stimulus from the sets of pairs. The stimulus changes after six correct responses. We hypothesize that the 930 mg/d group will exhibit greater attentional flexibility, as indicated by committing few errors.

### Exploratory Aim, Mental Health

We will assess our mental health exploratory objective with the ASEBA-web Youth Self Report.

<u>The Youth Self Report</u> (YSR) questionnaire for adolescents aged 11 to 18 years old assesses emotional and behavioral patterns. The participant reads 112 statements that describe children in their age group and rates how much each statement feels true for them on a 3-point Likert scale (not true, somewhat or sometimes true, very or often true). We hypothesize that the 930 mg/d group will have smaller raw scores of self-reported mental health problems on the ASEBA Youth Self Report, including smaller scores on a scale of internalizing behaviors (comprised of Anxious/Depressed, Withdrawn/Depressed, and Somatic Complaints) and smaller scores on a scale of externalizing behaviors (comprised of Rule-Breaking Behavior, Aggressive Behavior)

### Protocol Aim

This protocol aims to provide standardized instructions for administration of both the CANTAB and the ASEBA instruments in a virtually proctored testing environment to increase the feasibility of obtaining precise and valid measurements of adolescent cognition and mental health through remote testing.

### Testing Environment and Equipment Setup

During the informed consent call, the adolescent and their parent work with the study team to identify a suitable distraction-free environment for testing. This process includes identification of a quiet room and of a clutter-free desk or table at which the adolescent can complete testing.

After the informed consent call but before the first day of testing, we sent the study participant the necessary materials, including a study-standardized iPad, iPad case, and noise-canceling headphones. Providing standardized study materials to all participants is intended to prevent variation in test delivery that could arise from device hardware or software differences.

Before commencing each day of testing, the study proctor verifies that the participant’s cell phone is silenced and stored out of sight. Then, the proctor ensures the participant is in their pre-specified location, wearing study-provided noise-canceling headphones and is using the study-provided iPad.

### Testing Procedures

Participants will complete testing over three days within a one-week window, as depicted in Figure 1.

**Figure 1:**
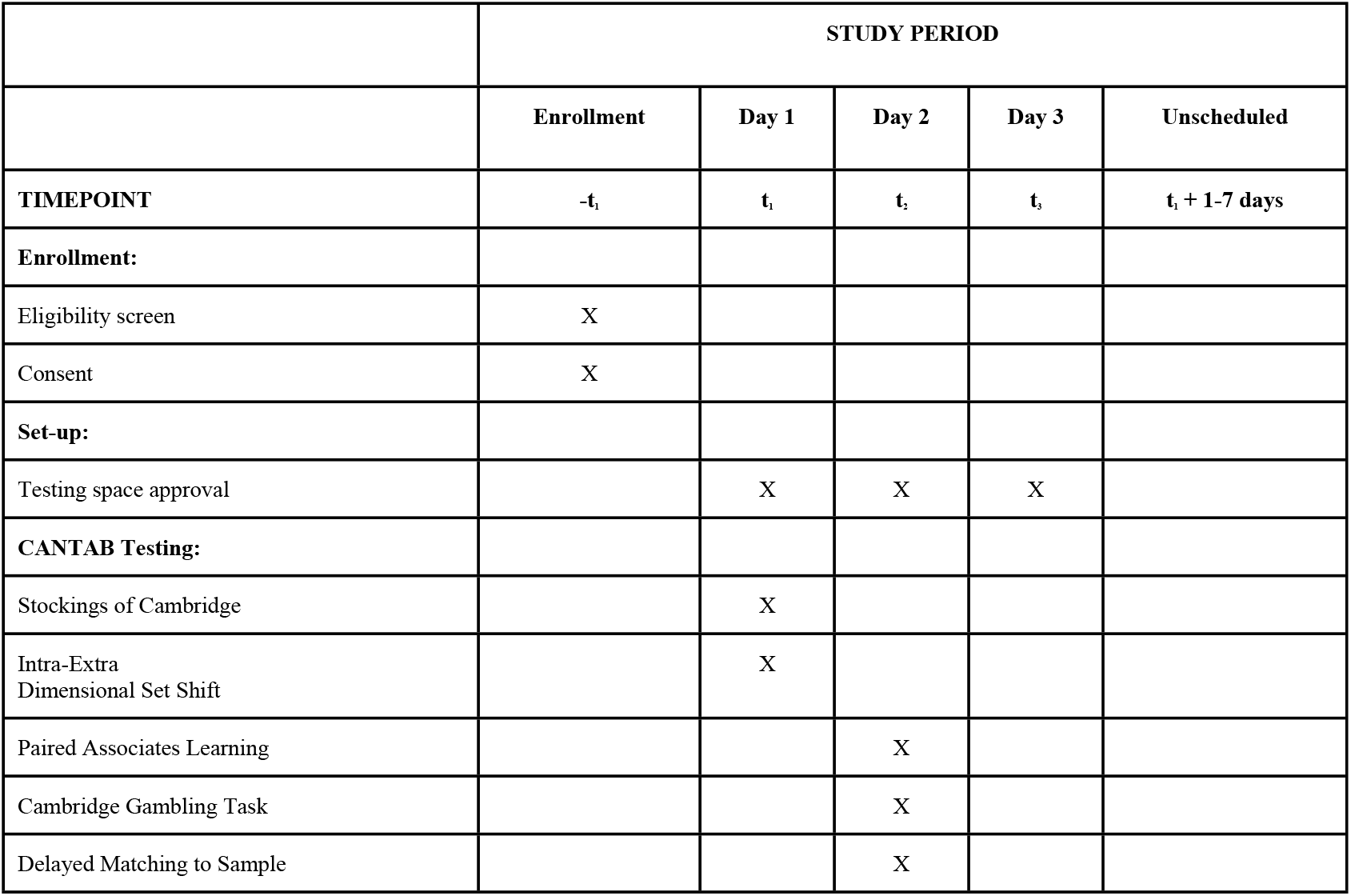
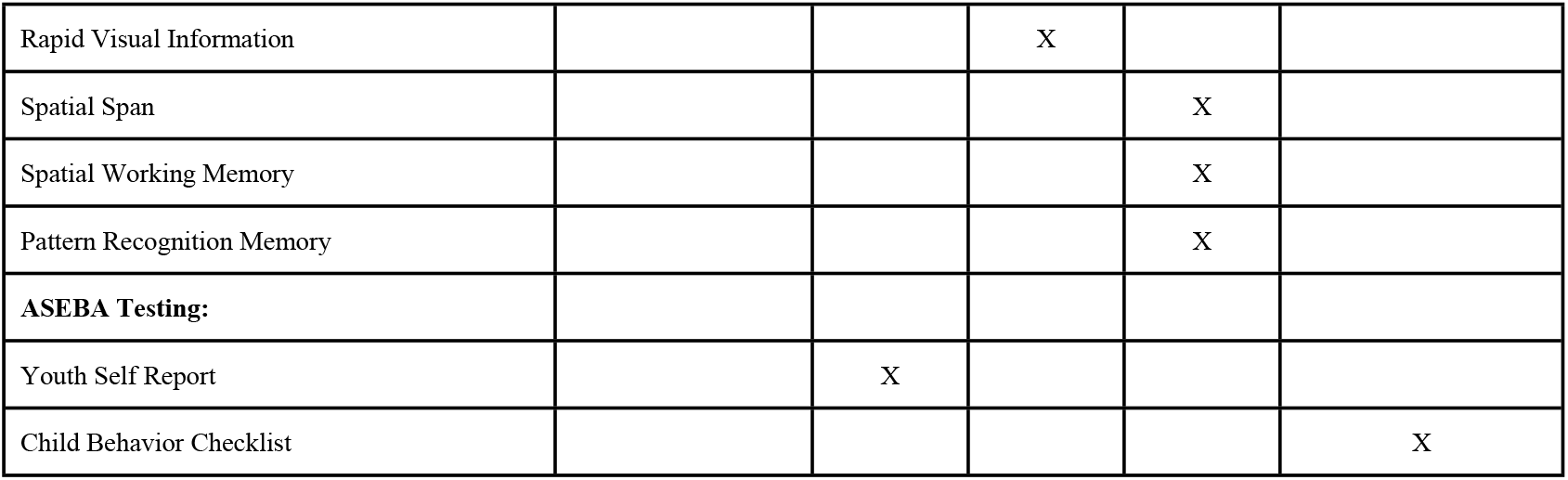
Participant schedule

### Day 1

When the participant joins the secure video-conferencing call, the study proctor guides the participant to use the iPad camera to confirm that the participant is in the pre-specified quiet testing space. After the proctor approves the testing environment, the participant returns to their seat and shares their screen for test proctoring. If testing space modifications are necessary, the proctor directs the participant to remove distractions or relocate.

The Day 1 protocol includes testing with the CANTAB and ASEBA software and requires approximately 37 minutes to complete. The participant receives an email from the proctor that includes links to their CANTAB and ASEBA tests. Upon the proctor’s instruction, the participant accesses the specified link(s). Both testing platforms provide simple onscreen and verbal instructions for each task, and the proctor can verbally clarify any potential participant questions. Participants complete testing in the following order: SOC, YSR, IED. The proctor offers an optional 3-minute break after the YSR. At the end of Day 1 testing, the proctor concludes the session with a reminder to the participant not to open the next CANTAB test link until the following testing session.

### Day 2

The proctor starts Day 2 testing with a re-assessment of the participant’s testing environment and verbal directions to access the CANTAB testing. The testing for Day 2 takes approximately 37 minutes to complete and only includes tests from the CANTAB software. The participants complete the testing in the following order: PAL, CGT, DMS, RVP. The proctor offers an optional 3-minute break after the CGT.

### Day 3

The testing on Day 3 takes approximately 35 minutes to complete and consists only of tests from the CANTAB. The participants complete the testing in the following order: SSP, SWM, PRM. A required break of 20 minutes occurs in the middle of the Pattern Recognition Memory test administration. At this time, the participant can get water, use the bathroom, and/or stretch.

### Data analysis plan Sample Size

The original controlled feeding study was designed to detect group differences in plasma choline levels. We estimated that a sample size of 26 (13 per treatment group) would achieve 80% power to detect a moderate effect size at an α of 0.05 (27). Our previous ancillary follow-up studies of offspring cognition, conducted during infancy and at age 7 years, demonstrated that (1) a sample size of 24 (12 per group) was sufficient to detected a statistically significant effect of the 930 mg/d prenatal choline intervention on infant processing speed and (2) that a sample size of 20 (11 and 9 participants from the 930/480 mg choline/d groups, respectively) was sufficient to obtain statistically significant group differences on several cognitive outcomes at age 7 years (25,28).

Based on these findings, we predict that the current sample size is sufficient to detect group differences at 14 years of age, should those differences exist.

### Descriptive Statistics for Sample Characteristics

We will present descriptive statistics of each group’s characteristics in tables which will include participant sex, level of education, parental level of education, native language, family income, head injury absence/presence, and any current medications and supplements. We will summarize continuous variables with means and standard deviations and categorical variables with absolute counts and relative frequencies. We will also provide a flow chart detailing participant attrition and missing data.

### Inferential Statistical Analysis

Acknowledging the risks inherent in using multiple endpoints in a small sample, we created an *a priori* analysis plan. Our specific aims focus on areas of cognitive functioning shown to be affected by maternal choline intake in prior animal and human studies. Below, we detail which variables we will use to assess each aim. For every outcome measure of interest, we will use linear models with a main effect of group and sex.

### Monitoring and data management

We store all identifiable data on password-protected servers and computers in locked offices. We will not disclose participants’ identities to anyone outside of the research team. Within the study team, we will share any identifiable data via a password-protected research server. We will assign participants numeric identifiers and will complete all data analysis with the participant number identifiers and not with any personally identifying information.

### Safety considerations

There are no direct significant risks associated with participation in this study. However, due to the sensitive nature of the YSR questions, we created a safety plan. If a participant, through their responses, discloses behavior or thoughts that raise concerns about their immediate safety or well-being, we will contact the parent(s) of the participant through a secure, password-protected communication platform. If deemed necessary, the study coordinator will recommend that the participant seek clinical evaluation.

## Ethical Approval

Ethical approval for the follow-up study was obtained from the Institutional Review Board for Human Participants at Cornell University in Ithaca, NY (USA). Written parental consent and written child assent from all participants and their parents was obtained by study coordinators before commencing any study activity.

## Discussion

This study is the first long-term follow-up of an RCT that assesses the enduring impact of prenatal choline intake on child cognition and mental health under conditions in which maternal choline intake is precisely known. The previous assessments of this cohort during infancy and at age 7 years revealed significant differences between the two groups in areas of information processing speed, working memory, and sustained attention (25,28). The present study follows these same children into adolescence, an age when group differences in cognition may be more indicative of lifelong effects of the early choline intervention (25,29). If our results support our hypotheses – that the higher choline group performs better than the lower choline group – they would provide the clearest evidence to date that the low choline intake of most pregnant women places their children at risk of subtle impairments in cognitive functioning.

Demonstrating that maternal choline intake has lasting effects on offspring cognition in humans, as it does in animals, would suggest that the many other long-lasting benefits of maternal choline supplementation reported in animals may also apply to humans. Specifically, in animals, maternal choline supplementation offers protection from some of the deleterious effects of perinatal exposure to neurotoxicants (e.g., alcohol, manganese) and lessens cognitive dysfunction in diverse neuropathological conditions including aging, Down syndrome, autism, and Alzheimer’s disease (2,3,8). Although the neuroprotective effect of maternal choline supplementation has received little attention in human studies, a few studies have evaluated this hypothesis in prenatal alcohol exposure, with promising results (11,33). If the results of our follow-up study indicate that the animal data translate to humans concerning lasting offspring cognitive effects, they open the exciting possibility that efforts to increase choline intake during pregnancy may not only improve cognitive functioning throughout the lifespan but also lessen the risk of cognitive impairments due to diverse environmental and genetic cognitive disorders (2,3,8).

Lastly, the use of standardized virtual cognitive testing offers an experimental technique that may be beneficial to many research areas. Particularly in the case of long-term cohort follow-up, such as the present study, wherein the participants may no longer reside near the research institution, it can be costly for participants to return. Thus, standardizing procedures for valid and reliable remote cognitive testing can be extremely valuable (34). However, in such studies, it is critical that the testing be closely proctored to avoid or minimize the possibility of inaccurate data due to the participants misunderstanding the instructions, losing focus in the tasks, or experiencing some other type of disruption in the testing. Standardized and proctored testing procedures ensure that the testing environment is conducive to acquiring high-quality data. The protocol developed for the present study establishes these procedures, ensures inter-proctor reliability in our study, and creates a precedent for use by future scientists interested in virtual cognitive testing. The ability to collect data in a virtual setting affords the opportunity to reach study populations that are distant from the research institution, potentially increasing the generalizability of study findings through the inclusion of more diverse participants (35).

## Conclusion

This study aims to determine the long-term impacts of prenatal choline intake on adolescent offspring’s cognitive functioning and mental health. Leveraging a rigorous double-blind controlled feeding trial that achieved precise control over the participants’ total daily choline intakes, we have a unique opportunity to assess the long-term effects of prenatal choline supplementation. Our findings may offer pivotal insights into the developmental trajectories associated with prenatal choline intake, potentially influencing dietary recommendations and prenatal care practices. Furthermore, the virtual testing framework in this study ensures the feasibility and reliability of long-term cognitive assessments and sets a precedent for future research methodologies, increasing accessibility and participation in cognitive studies.

Ultimately, this research may underscore the importance of prenatal choline intake for lifelong cognitive and mental health outcomes in offspring.

## Data Availability

No datasets have yet been generated or analysed during the current study. All relevant data from this study will be made available upon study completion.

## References

1. Zeisel SH. Choline: Critical Role During Fetal Development and Dietary Requirements in Adults [Internet]. Available from: http://www.nal.usda.gov/fnic/foodcomp/Data/Choline/Choline.html

2. McCann JC, Hudes M, Ames BN. An overview of evidence for a causal relationship between dietary availability of choline during development and cognitive function in offspring. Vol. 30, Neuroscience and Biobehavioral Reviews. 2006. p. 696–712.

3. Meck WH, Williams CL. Metabolic imprinting of choline by its availability during gestation: Implications for memory and attentional processing across the lifespan. In: Neuroscience and Biobehavioral Reviews. Elsevier Ltd; 2003. p. 385–99.

4. Meck WH, Williams CL. Choline supplementation during prenatal development reduces proactive interference in spatial memory [Internet]. Vol. 118, Developmental Brain Research. 1999. Available from: www.elsevier.comrlocaterbres

5. Williams CL, Meck WH, Dee Heyer D, Loy R. Hypertrophy of basal forebrain neurons and enhanced visuospatial memory in perinatally choline-supplemented rats. Vol. 794, Brain Research. 1998.

6. Cheng RK, MacDonald CJ, Williams CL, Meck WH. Prenatal choline supplementation alters the timing, emotion, and memory performance (TEMP) of adult male and female rats as indexed by differential reinforcement of low-rate schedule behavior. Learning and Memory. 2008 Mar;15(3):153–62.

7. Yang Y, Liu Z, Cermak JM, Tandon P, Sarkisian MR, Stafstrom CE, et al. Protective Effects of Prenatal Choline Supplementation on Seizure-Induced Memory Impairment [Internet]. 2000. Available from: http://www.jneurosci.org/cgi/content/full/4733

8. Moon J, Chen M, Gandhy SU, Strawderman M, Levitsky DA, Maclean KN, et al. Perinatal choline supplementation improves cognitive functioning and emotion regulation in the Ts65Dn mouse model of down syndrome. Behavioral Neuroscience. 2010;124(3):346–61.

9. Strupp BJ, Powers BE, Velazquez R, Ash JA, Kelley CM, Alldred MJ, et al. Maternal choline supplementation: A potential prenatal treatment for Down syndrome and Alzheimer’s disease HHS Public Access Author manuscript. Vol. 10962, Curr Alzheimer Res. Nathan Kline Institute; 2016.

10. Ryan SH, Williams JK, Thomas JD. Choline supplementation attenuates learning deficits associated with neonatal alcohol exposure in the rat: Effects of varying the timing of choline administration. Brain Res [Internet]. 2008 Oct 10 [cited 2024 Aug 8];1237:91. Available from: /pmc/articles/PMC2646103/

11. Jacobson SW, Carter RC, Molteno CD, Stanton ME, Herbert JS, Lindinger NM, et al. Efficacy of Maternal Choline Supplementation During Pregnancy in Mitigating Adverse Effects of Prenatal Alcohol Exposure on Growth and Cognitive Function: A Randomized, Double-Blind, Placebo-Controlled Clinical Trial. Alcohol Clin Exp Res [Internet]. 2018 Jul 1 [cited 2024 Aug 1];42(7):1327–41. Available from: https://pubmed.ncbi.nlm.nih.gov/29750367/

12. Fan G, Feng C, Wu F, Ye W, Lin F, Wang C, et al. Methionine choline reverses lead-induced cognitive and N-methyl-d-aspartate receptor subunit 1 deficits. Toxicology. 2010 Jun 4;272(1–3):23–31.

13. Latta DM, Donaldson WE. Lead toxicity in chicks: interactions with dietary methionine and choline. J Nutr [Internet]. 1986 [cited 2025 Jan 12];116(8):1561–8. Available from: https://pubmed.ncbi.nlm.nih.gov/3761012/

14. Howard SL, Beaudin SA, Strupp BJ, Smith DR. Maternal choline supplementation lessens the behavioral dysfunction produced by developmental manganese exposure in a rodent model of ADHD. Neurotoxicol Teratol. 2024;102:107337.

15. Glenn MJ, Adams RS, McClurg L. Supplemental dietary choline during development exerts antidepressant-like effects in adult female rats. Brain Res [Internet]. 2012 Mar 14 [cited 2024 Dec 16];1443:52. Available from: https://pmc.ncbi.nlm.nih.gov/articles/PMC3327365/

16. Schulz KM, Pearson JN, Gasparrini ME, Brooks KF, Drake-Frazier C, Zajkowski ME, et al. Dietary choline supplementation to dams during pregnancy and lactation mitigates the effects of in utero stress exposure on adult anxiety-related behaviors. Behavioural brain research [Internet]. 2014 Jul 15 [cited 2024 Aug 1];268:104–10. Available from: https://pubmed.ncbi.nlm.nih.gov/24675162/

17. Boeke CE, Gillman MW, Hughes MD, Rifas-Shiman SL, Villamor E, Oken E. Choline intake during pregnancy and child cognition at age 7 years. Am J Epidemiol. 2013 Jun 15;177(12):1338–47.

18. Wu BTF, Dyer RA, King DJ, Richardson KJ, Innis SM. Early second trimester maternal plasma choline and betaine are related to measures of early cognitive development in term infants. PLoS One. 2012;7(8):e43448.

19. Hunter SK, Hoffman MC, McCarthy L, D’Alessandro A, Wyrwa A, Noonan K, et al. Black American Maternal Prenatal Choline, Offspring Gestational Age at Birth, and Developmental Predisposition to Mental Illness. Schizophr Bull. 2021 Jul 8;47(4):896–905.

20. Signore C, Ueland PM, Troendle J, Mills JL. Choline concentrations in human maternal and cord blood and intelligence at 5 y of age. Am J Clin Nutr. 2008 Apr;87(4):896–902.

21. Hunter SK, Hoffman MC, D’Alessandro A, Walker VK, Balser M, Noonan K, et al. Maternal prenatal choline and inflammation effects on 4-year-olds’ performance on the Wechsler Preschool and Primary Scale of Intelligence-IV. J Psychiatr Res [Internet]. 2021 Sep 1 [cited 2024 Aug 1];141:50–6. Available from: https://pubmed.ncbi.nlm.nih.gov/34174557/

22. Hunter SK, Hoffman MC, D’Alessandro A, Wyrwa A, Noonan K, Zeisel SH, et al. Prenatal choline, cannabis, and infection, and their association with offspring development of attention and social problems through 4 years of age. Psychol Med. 2022 Oct;52(14):3019–28.

23. Freedman R, Hunter SK, Law AJ, Wagner BD, D’Alessandro A, Christians U, et al. Higher Gestational Choline Levels in Maternal Infection Are Protective for Infant Brain Development. J Pediatr. 2019 May;208:198-206.e2.

24. Cheatham CL, Goldman BD, Fischer LM, Da Costa KA, Reznick JS, Zeisel SH. Phosphatidylcholine supplementation in pregnant women consuming moderate-choline diets does not enhance infant cognitive function: A randomized, double-blind, placebo-controlled trial. American Journal of Clinical Nutrition. 2012 Dec 1;96(6):1465–72.

25. Caudill MA, Strupp BJ, Muscalu L, Nevins JEH, Canfield RL. Maternal choline supplementation during the third trimester of pregnancy improves infant information processing speed: A randomized, double-blind, controlled feeding study. FASEB Journal. 2018 Apr 1;32(4):2172–80.

26. Ross RG, Hunter SK, McCarthy L, Beuler J, Hutchison AK, Wagner BD, et al. Perinatal Choline Effects on Neonatal Pathophysiology Related to Later Schizophrenia Risk. American Journal of Psychiatry. 2013 Mar;170(3):290–8.

27. Yan J, Jiang X, West AA, Perry CA, Malysheva O V., Devapatla S, et al. Maternal choline intake modulates maternal and fetal biomarkers of choline metabolism in humans. Am J Clin Nutr [Internet]. 2012 May 1 [cited 2025 Jan 12];95(5):1060–71. Available from: https://pubmed.ncbi.nlm.nih.gov/22418088/

28. Bahnfleth CL, Strupp BJ, Caudill MA, Canfield RL. Prenatal choline supplementation improves child sustained attention: A 7-year follow-up of a randomized controlled feeding trial. FASEB Journal. 2022 Jan 1;36(1).

29. Nyongesa MK, Ssewanyana D, Mutua AM, Chongwo E, Scerif G, Newton CRJC, et al. Assessing executive function in adolescence: A scoping review of existing measures and their psychometric robustness. Vol. 10, Frontiers in Psychology. Frontiers Media S.A.; 2019.

30. Institute of Medicine (US) Standing Committee on the Scientific Evaluation of Dietary Reference Intakes and its Panel on Folate OBV and C. The B Vitamins and Choline: Overview and Methods. 1998 [cited 2024 May 22]; Available from: https://www.ncbi.nlm.nih.gov/books/NBK114324/

31. Soares FC, de Oliveira TCG, de Macedo LD e. D, Tomás AM, Picanço-Diniz DLW, Bento-Torres J, et al. CANTAB object recognition and language tests to detect aging cognitive decline: An exploratory comparative study. Clin Interv Aging. 2014 Dec 19;10:37–48.

32. MacMillan NA. Signal Detection Theory. In: Stevens’ Handbook of Experimental Psychology. Wiley; 2002.

33. Warton FL, Molteno CD, Warton CMR, Wintermark P, Lindinger NM, Dodge NC, et al. Maternal choline supplementation mitigates alcohol exposure effects on neonatal brain volumes. Alcohol Clin Exp Res. 2021 Sep;45(9):1762–74.

34. Backx R, Skirrow C, Dente P, Barnett JH, Cormack FK. Comparing web-based and lab-based cognitive assessment using the cambridge neuropsychological test automated battery: A within-subjects counterbalanced study. J Med Internet Res. 2020 Aug 1;22(8).

35. Hildenbrand L, Wiley J. Mental counters as an online tool for assessing working memory capacity. Behav Res Methods. 2023;

